# ‘It helps my anxiety because I’m managing my breathlessness’: a qualitative exploration of anxiety and breathlessness in patients with advanced chronic respiratory disease receiving specialist palliative care

**DOI:** 10.1101/2025.09.03.25334785

**Authors:** Lucy Bleazard, Kate Walker, Sue Ashton, Christina Faull

**Author notes:** Corresponding author., LOROS Centre for Excellence, Groby Road, Leicester, LE3 9QE.

## Abstract

**Objectives:** To explore the lived experiences of anxiety related to breathlessness in patients who are receiving specialist palliative care for advanced chronic respiratory disease (CRD), such as COPD and interstitial lung diseases.

**Methods:** This qualitative exploration formed part of a mixed-methods feasibility study of a novel intervention. Participants receiving specialist palliative care for CRD at two UK hospice sites engaged in semi-structured interviews. Data were analysed using thematic analysis.

**Results:** Three key organising themes emerged: 1) Understanding my breathlessness – participants described their breathlessness as progressive, frightening, and restrictive. 2) Understanding my anxiety – described as emotional distress with a profound physical component, often compounded by external life stressors. 3) Vicious circle interlinks breathlessness and anxiety – a circular bi-directional relationship where anxiety could not be separated from breathlessness. Participants often employed non-pharmacological strategies to manage both their anxiety and breathlessness interdependently.

**Significance of Results:** Our findings demonstrate the interdependency between anxiety and breathlessness for patients with a range of CRD diagnoses. The shared symptom experience across a range of CRD supports the value of unified management approaches to these two symptoms in specialist palliative care breathlessness clinics.

## Introduction

Chronic respiratory disease (CRD) is one of the leading causes of death worldwide (Adeloye *et al*. 2022; Momtazmanesh *et al*. 2023). Encompassing a range of conditions including chronic obstructive pulmonary disease (COPD) and interstitial lung diseases (ILD), CRD is characterised by persistent, progressive breathlessness (*GOLD*, 2025; Maher 2024; Müllerová *et al*. 2014). This debilitating physical symptom is often the primary focus of symptom management, and whilst this is both necessary and justified, many patients with CRD will experience substantial psychological distress which is frequently under-recognised and insufficiently addressed (Rzadkiewicz *et al*. 2016).

Anxiety constitutes a significant component of this psychological burden. Up to a third of patients with COPD will meet the criteria for diagnosis with generalised anxiety disorder (GAD) (Willgoss and Yohannes 2013), with comparable estimates for interstitial lung diseases (Yohannes 2020). The relationship between anxiety and the physiological experience of breathlessness in CRD is complex, as feelings of anxiety amplify the perception of breathlessness and vice-versa (Bailey 2004). As a result, the management of anxiety related to breathlessness in advanced CRD presents a significant clinical challenge.

Specialist palliative care (SPC) services, particularly holistic multi-disciplinary breathlessness clinics, have demonstrated efficacy in supporting patients to manage their breathlessness (Higginson *et al*. 2014; Farquhar *et al*. 2016). However there has been little exploration of how patients utilising these services experience and make sense of their anxiety related to their breathlessness. This paper explores these lived experiences for patients utilising SPC for a range of CRD diagnoses.

This work forms part of a wider feasibility study exploring the potential use of neuromodulation in the management of anxiety related to breathlessness.

## Methods

This work is part of a mixed-methods interventional feasibility study over 12 weeks included a single semi-structured interview with participants at the end of their involvement in the study. We report here analysis of those interviews with respect to participants experiences of anxiety related to breathlessness. Participants were recruited at two specialist palliative care services, both of which provide multidisciplinary outpatient services for patients with advanced CRD.

### Participants

Participants were receiving SPC at the recruiting hospice site for CRD of any aetiology. Diagnoses were recorded as documented in their electronic patient record. Eligibility criteria included an IPOS (Hearn and Higginson 1999) score of ≥2 for both ‘shortness of breath’ and ‘anxious/worried’ domains, which had been routinely administered as part of clinical care. All patients attending SPC outpatient breathlessness services were screened for eligibility, and a purposive sampling strategy was used to ensure a representation across a range of CRD.

### Recruitment

Members of the patients’ usual clinical team introduced the study to those identified through screening. Individuals who expressed an interest in the study were provided with a written participant information sheet and contacted by a member of the research team to confirm eligibility and willingness to proceed. Written informed consent was obtained from eligible patients prior to enrolment.

### Data collection

Participants were invited to participate in an in-depth semi-structured telephone interview with an experienced qualitative researcher towards the end of the 12-week study period to explore their lived experiences of anxiety and breathlessness. There was no prior established relationship between the researcher conducting the interviews and the participants. Interviews were audio-recorded onto an encrypted recorder. Audio files were transcribed verbatim and anonymised at the point of transcription.

### Data analysis

Interview data were analysed using thematic analysis as a means of organising the data into patterns and themes (Braun and Clarke 2006, 2021) and to highlight differences and similarities between participants’ accounts (Creswell 2009). Thematic analysis allows researchers to take a structured approach to managing and analysing the data, and so is suitable for identifying patterns across and within the interviews in relation to the attitudes, beliefs, perceptions and experiences of the participants (Clarke and Braun 2017; Creswell 2009). Themes were developed using an inductive approach at the semantic level, therefore based on the explicit or surface meanings of the data (Braun and Clarke 2021). Data analysis followed the six-step framework that is consistently used for thematic analysis (Braun and Clarke 2006, 2021).

Verbatim (anonymised) quotes were identified and included to promote verifiability and trustworthiness of the analysis (Silverman 2013). To ensure that the findings were the experiences of those interviewed, strategies advocated by Shenton (2004) were implemented to enable credibility and confirmability.

### Ethical approvals

Ethical approval for the study was gained through NHS HRA REC (reference 23/LO/0276).

## Results

Eleven participants completed an interview. Participant characteristics are shown ***Table 1***.

**Table 1.**
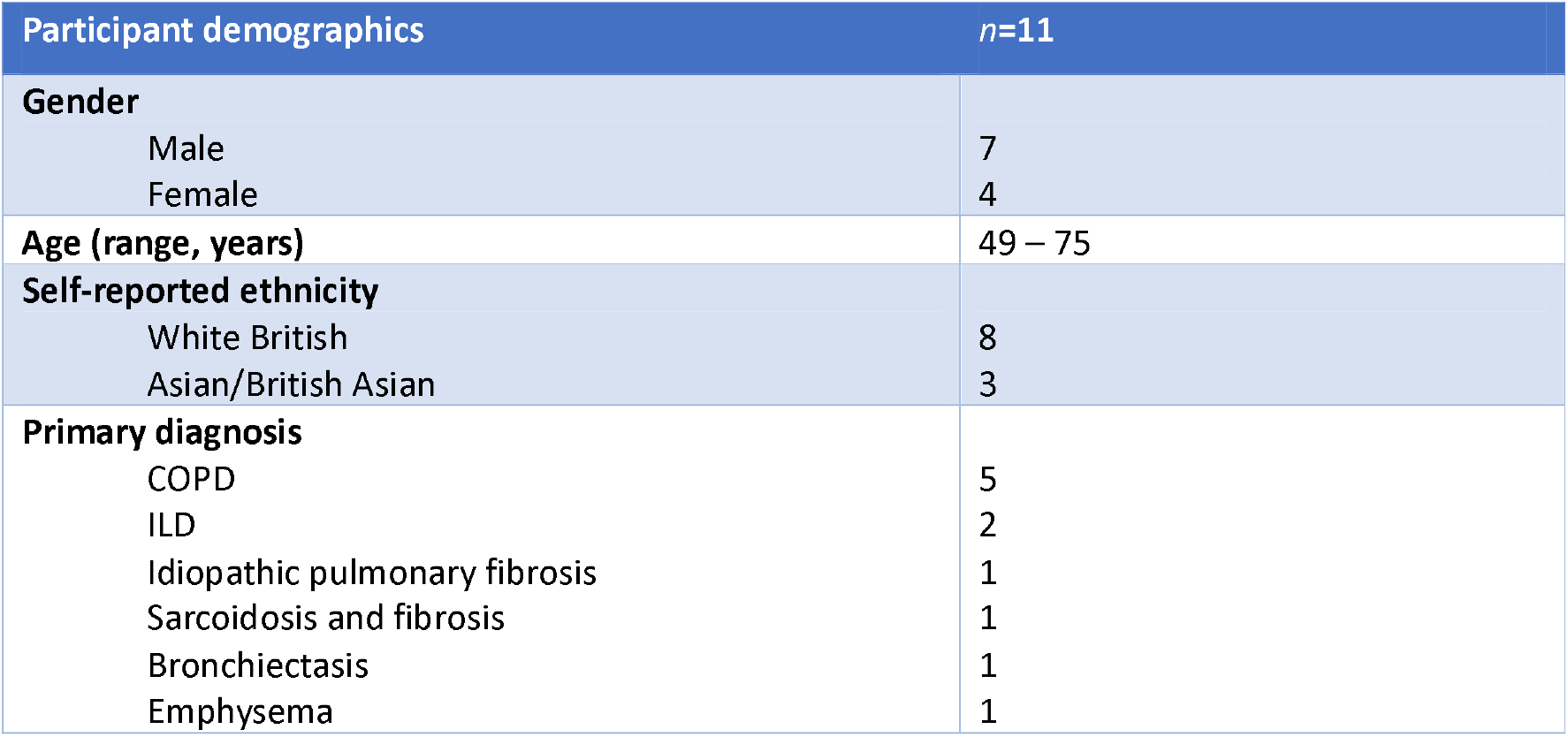
Participant demographics.

| Participant demographics | *n*=11 |
| --- | --- |
| **Gender** |  |
| Male | 7 |
| Female | 4 |
| **Age (range, years)** | 49 – 75 |
| **Self-reported ethnicity** |  |
| White British | 8 |
| Asian/British Asian | 3 |
| **Primary diagnosis** |  |
| COPD | 5 |
| ILD | 2 |
| Idiopathic pulmonary fibrosis | 1 |
| Sarcoidosis and fibrosis | 1 |
| Bronchiectasis | 1 |
| Emphysema | 1 |

Three organising themes were developed, presented in *Figure 1*: (1) Understanding my breathlessness; (2) Understanding my anxiety; and (3) Vicious circle interlinks breathlessness and anxiety. The first two organising themes are represented by several subthemes, while the third organising theme was developed as a standalone theme.

**Figure 1:**
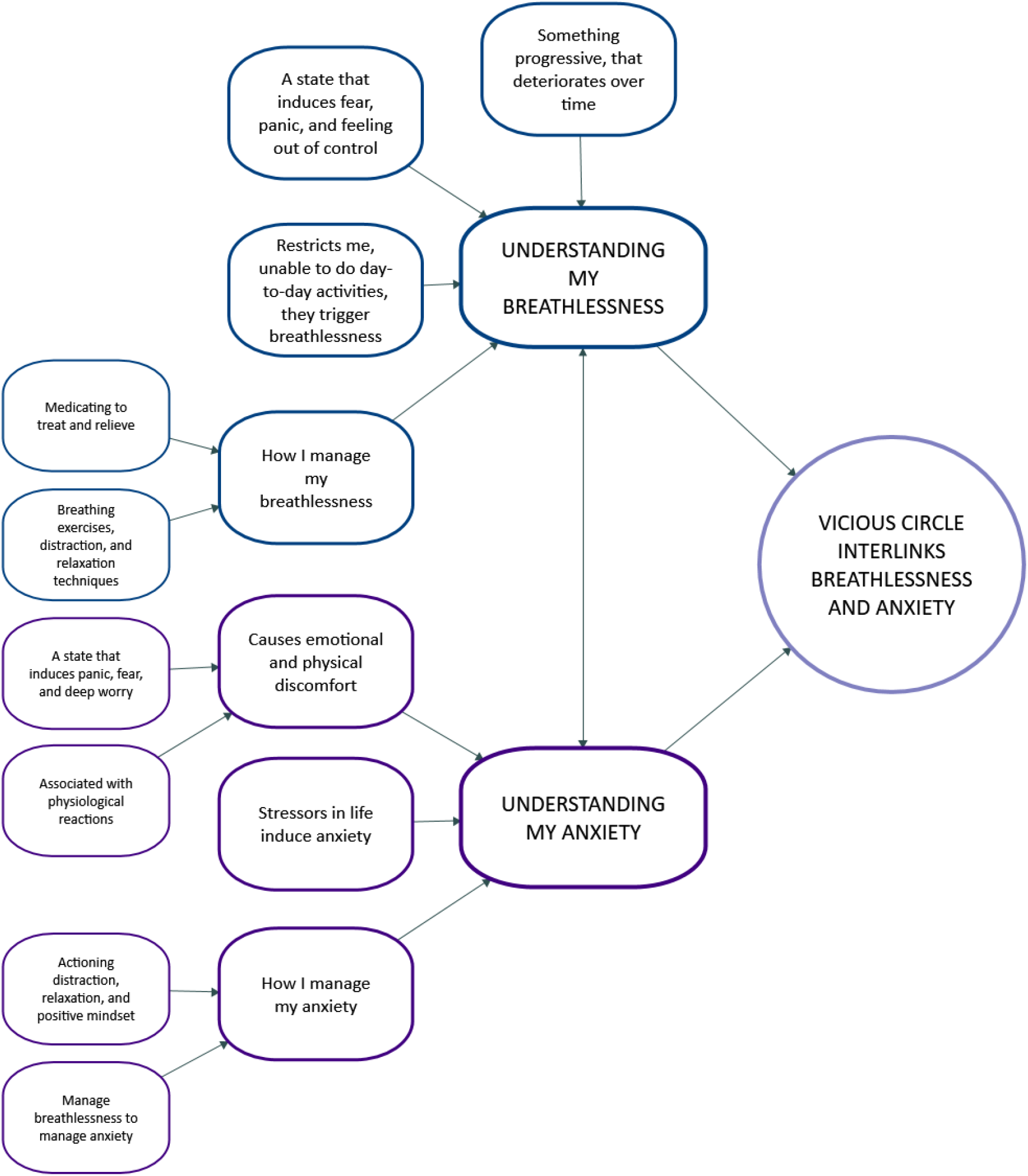
Themes and subthemes of patients' experience of anxiety and breathlessness.

### Understanding my breathlessness

This organising theme concerns participants’ experience and understanding of their breathlessness; what it means to each person, how it feels, the impact on their daily lives, and their management strategies. This organising theme is represented by four individual themes.

#### Something progressive, that deteriorates over time

Participants revealed how their breathlessness caused them to realise they were living with a progressive illness, that has and would continue to worsen over time.

> **P5**: Well, it started off, I wouldn’t say it was mild, it was enough for me to notice it. So, throughout those 10 years, it has just slowly gotten worse. So now it is probably the worst it’s always, it’s ever been. But it, as time’s getting on, it is getting worse and worse.

Within the narratives, there was reference by several to this being something that was ‘*deteriorating*’ over time and as such was preventing them from doing things they previously were able to do.

> **P7**: Deterioration in my condition. It is simply the deterioration in my condition. I could go out shopping on my own, just with my oxygen. I was driving. I would go out on my own as well. Whereas now, I can’t do that anymore. It’s the, the speed of it deteriorating.

This was accompanied by participants acknowledging their disease was never going to improve, and that it can only be managed rather than cured.

> **P6**: There is nothing that will improve it, but I’ve just got to try and manage it. It made me realise just how bad things could be. Now looking at it, well I am breathless all day, you know, I am never going to get any better.

#### A state that induces fear, panic, and feeling out of control

All participants related to the fact that their breathlessness was associated with extreme feelings of fear and panic, even ‘*terror*’. Many described it as being distinctly different to any sensation they had experienced before.

> **P2**: It’s scary. Sometimes you feel that this is your last breath.

Some participants offered illustrative analogies to express this fear and panic, and in doing so offered a rich description of what this experience is really like for them.

> **P1**: Scary. It’s like trying to climb a mountain with a plastic bag over your head.
>
> **P7**: So, it’s like permanently having a bag over your head or a scarf over your mouth and nose.
>
> **P11**: So that feeling of your drowning almost but drowning in air.

In addition to this, participants also expressed how this fear and panic was experienced as physiological reactions. Examples included becoming hot, an increased heart rate, and palpitations.

> **P8**: It is when I get breathless, then heart starts racing, then I start panicking. It’s like I am really burning hot, like I am on fire.

#### Restricts me, unable to do day-to-day activities, they trigger breathlessness

All participants identified that doing certain activities triggers their breathlessness. Exertion of any type was felt to be a trigger, even something that may be perceived as very low-intensity activity. Many found climbing the stairs becoming a particular difficulty.

> **P5**: If I exert myself in any way, the stairs … even now I’m talking to you, I’m getting slightly out of breath. So walking, any physical activity, bending down, anything at all causes me to become very breathless.

This all restricts the individual and means they are unable to engage in their usual activities of daily living. Participants reflected that these were activities most people would take for granted. These included washing and dressing, playing with their children, and housework:

> **P5**: Playing with your children, that probably had the most impact on me, it makes you realise that yes, this is actually impacting a lot of my lifestyle now, practically all of it.

#### How I manage my breathlessness

Participants manage their breathlessness in a variety of ways. This emerges through two subthemes, one relating to pharmacological management and one to non-pharmacological strategies.

### Medicating to treat and relieve

Many participants use pharmacological management to control their breathlessness. Regular medication, primarily a variety of inhaled medication and oral mucolytics, were described by most participants. This was perceived as controlling their underlying disease, and therefore managing their breathlessness.

> **P8**: I have regular medication - inhalers, I have tablets as well. With the tablets they are for like the phlegm and things like that. The only thing I have got for the COPD is the Trimbow inhaler, that is the only medication there really is for it.

The other use of medication was to relieve their breathlessness at times of escalation, by taking as-required inhalers or oral opioids. One participant described using supplemental oxygen to control their breathlessness when it became overwhelming.

> **P3**: Taking oxygen obviously makes it [episode of breathlessness] better. Nights can be a bit of a problem… I just take some morphine or pull on me inhaler or something like that. I take the Ventolin whenever I need it.

### Breathing exercises, distraction, and relaxation techniques

Many participants found the non-pharmacological techniques they had been taught helpful and employed these regularly, including mindfulness, grounding, and breathing exercises.

> **P1**: So, I do my mindfulness and grounding techniques. So, it’s the 5,4,3,2,1. I also do a lot of breathing exercises where I breathe in for four and exhale for five. I have a window that’s an oblong and I breathe in and out on the ups and downs and the sides of the window and stuff.
>
> Similarly, some participants also discussed that techniques they used were around relaxation, feeling calmer, or trying to sleep.

> **P2**: Relaxing. Definitely trying to relax. And have a sleep if you can. I have to have one every afternoon just for an hour, or even if you don’t go sleep, just go and lay on the bed.

Many participants employed distraction techniques to divert their attention from their breathlessness, including reading, watching television, or using their mobile phone.

> **P5**: I use a form of distraction, not be thinking about, you know, a) my cough and b) my breathing. So, I use a form of distraction. I always have my phone with me, so the main thing I’ll do just go on my phone and start reading the news, or go on Twitter or, or something, that will take my mind off, what’s actually happening to me.

### Understanding my anxiety

Similarly to our first organising theme, this concerns participants understanding of their anxiety in terms of how it feels, its meaning and impact on their lives, and their management strategies. It is represented by three individual themes.

#### Causes emotional and physical discomfort

Participants experience anxiety as a sensation of discomfort, both emotional and physical. This is presented as two subthemes to reflect each of these domains.

### A state that induces panic, fear, and deep worry

Anxiety comprises a number of unpleasant and uncomfortable emotions. Participants had different words for describing how anxiety made them feel, but commonly used terminology was panic:

> **P5**: It’s like a panic feeling, and I, it, I feel a bit zoned out as well. And I can’t concentrate on anything around me, and, yeah, so this, it’s the feeling panicked;
>
> fear:
>
> **P4**: The anxiety is like a feeling of fear, it’s like fear. I mean I went to the toilet one time, and I got off the toilet and just fell, I collapsed. That really got me, it really did honestly;
>
> and feeling scared:
>
> **P8**: It is like when you are anxious isn’t it. I go shaky and stuff. It is like I don’t know; I am just scared all the time.

### Associated with physiological reactions

Alongside the distressing emotions associated with their anxiety, participants also discussed having physical reaction to their anxiety. Examples participants described included feeling their heart rate increasing:

> **P2**: Anxiety is when your heart rate going. And you sort of hyperventilated. Well, I can feel my heartbeat and it probably isn’t and it feels like it’s going fast you know;

sweating palms and feeling hot:

> **P5**: I feel panicked a little bit, and my heart starts raising. I, my palms start sweating. I feel hot. Those are the physical feelings that I get;

and nausea:

> **P6**: Yes, it is dread. It is fear. You get anxious. You, it makes me, it even makes me feel sick and I start heaving and I’m not being sick. It really makes me start to retch like.

#### Stressors in life induce anxiety

Participants described having a baseline ‘general anxiety’ because of their diagnosis, prognosis, and knowledge of their disease. Different stressors would then occur during day-to-day life that would build on and worsen their baseline anxiety.

**P11**: I don’t know what came first, the chicken or the egg. I, I’m not sure. My anxiety is tied to a degree to my diagnosis. It’s tied to a lot of other things other than my breathlessness.

A wide range of events were perceived as being stressors, from taking their child to college to family bereavements. The time of year was frequently cited, with the arrival of winter or the Christmas period often worsening their feelings of anxiety.

> **P1**: Again, you’ve caught me at the wrong time, with anxiety because I’ve had so much going on. It’s Christmas and you know, sort of being with family and everything.

#### How I manage my anxiety

This theme comprises two subthemes that are very much interlinked. The first relates to strategies implemented when participants are experiencing their ‘general activity’. The second relates to the management of their breathlessness when this is seen as the cause of their anxiety; thereby if breathlessness is managed, the associated anxiety will be too.

### Actioning distraction, relaxation, and positive mindset

When participants feel generally anxious, they implement strategies, tools and techniques such as distraction and relaxation. The strategies described were often similar to those used to manage exacerbations of breathlessness.

> **P2**: Just try to keep as busy as I can. Keep me mind occupied, either reading or games or television or something. Try not to think about it too much you know. Mind over matter I suppose.

Many participants described dealing with their anxiety by attempting to change their thinking, to adopt a positive mindset, or to create ‘positive energy’, which was notably absent when discussing how to manage their breathlessness.

> **P10**: It’s the thought process you know, all the negative thinking which create anxiety and I, try to build positive feelings all the time. I try to counter my activity with positive energy. I tried to feel positive energy within me.

### Manage breathlessness to manage anxiety

Participants often described dealing with their breathlessness as a method of dealing with their anxiety, acknowledging the link between the two. This leads us into our final organising theme.

> **P4**: I have just got to sit down and start breathing, slowly. So, because it’s related to my breathlessness, I manage my anxiety by managing my breathlessness. It takes the pressure off – like a boiling kettle! I guess it helps with my anxiety because I’m managing my breathlessness.

### Vicious circle interlinks breathlessness and anxiety

This final organising theme draws together the experiences of participants of their anxiety and their breathlessness, as many discuss how these are inextricably linked to one another. Participants experience breathlessness causing anxiety, but then anxiety causes breathlessness - a bi-directional causal relationship between anxiety and breathlessness, mediated by fear and panic of both experiences. Participants described this as part of their everyday life, not just in times of acute exacerbation.

> **P2**: I think it’s just terrifying when you can’t breathe, so it just forces anxiety….so I breathe, and I can’t really breathe, and I feel like I am suffocating. So, it’s definitely this anxiety comes when I’m actually feeling breathless, and then being anxious makes me more breathless.

As described by many participants, they experience this as a vicious circle. Participants feel that they keep going round in a continual circle of breathlessness-anxiety-breathlessness. Management techniques can help to reduce the sensations or potentially, albeit temporarily, break the circle.

> **P9**: Because you can’t breathe, you do get anxious, you get panicky. So, you’re breathless, so that makes you anxious, and then as you are anxious, that then makes you more breathless, as it were, it’s a vicious cycle.

## Discussion

This study offers valuable insight into the lived experiences of patients with a range of CRD receiving SPC, with a particular focus on anxiety and its complex relationship with breathlessness. Participants consistently reported a bi-directional interplay between these symptoms, often describing a ‘vicious circle’ in which the triggering of anxiety worsened breathlessness and vice versa. Bailey’s dyspnoea-anxiety-dyspnoea cycle (2004) during acute exacerbations of COPD is evident amongst our participants at baseline, beyond episodes of acute illness.

The relentless daily struggle with breathlessness was identified as one of many triggers for a deterioration in mental wellbeing. In addition to dyspnoea-related anxiety, some participants reported a generalised, constant anxiety that was not directly attributable to breathlessness. Notably, no participants had received a formal anxiety disorder diagnosis despite reporting symptoms consistent with GAD. This reflects the existing literature indicating patients’ psychological distress is highly prevalent yet often under-recognised and under-treated (Willgoss and Yohannes 2013; Yohannes 2020).

The experience of anxiety was primarily reported by description of the physical manifestations, and many found describing the underlying psychological dimensions of their anxiety more challenging. This suggests a difficulty in conceptualising anxiety and breathlessness as discrete symptoms. Rather, participants appeared to experience them as mutually reinforcing with significant overlap. This is consistent with current literature which emphasises the shared physical overlap between breathlessness and anxiety is a major contributor to worsening of both symptoms (Christiansen *et al*. 2023).

The similarities in symptom experience also extended to how participants employed self-management strategies. Non-pharmacological techniques were frequently described regardless of whether anxiety or breathlessness was perceived as the overwhelming symptom. Mindfulness, relaxation, breathing exercises, and distraction were often used to alleviate distress from both symptoms, and these approaches were perceived as accessible in times of deterioration. These techniques are a cornerstone of management in specialist palliative care breathlessness service (Farquhar *et al*. 2016; Powell 2014) and our findings support this holistic approach.

Participants’ descriptions of anxiety and management strategies were consistent regardless of their underlying respiratory diagnosis. These findings suggest that the symptom experience is comparable across the broad range of non-malignant respiratory conditions. This supports the suitability of a unified approach to care in specialist breathlessness clinics, irrespective of diagnosis.

### Strengths and limitations

This study encompasses patients with a range of CRD diagnoses rather than focusing on a single disease. All participants were identified as having high levels of anxiety and breathlessness at baseline using the IPOS, which is a widely used tool in the palliative care setting. The sample size is small yet reasonable for a qualitative study using thematic analysis, and thematic saturation was reached. The participants included in this analysis were recruited from two sites in a single region, perhaps limiting wider generalisability.

## Conclusion

This study provides valuable insight into the intricate relationship between anxiety and breathlessness in patients with a range of CRD accessing specialist palliative care services. Our findings emphasise that anxiety cannot be separated from breathlessness, and there is an ongoing need for holistic support for patients with anxiety related to breathlessness in non-malignant conditions.

## Data Availability

All data produced in the present work are contained in the manuscript.

## Acknowledgements

The authors would like to thank the participants of this study for so generously giving their time.

## References

Adeloye D, Song P, Zhu Y, Campbell H, Sheikh A and Rudan I (2022) Global, regional, and national prevalence of, and risk factors for, chronic obstructive pulmonary disease (COPD) in 2019: a systematic review and modelling analysis. 10(5), 447–458. 10.1016/S2213-2600(21)00511-7.

Bailey PH (2004) The dyspnea-anxiety-dyspnea cycle--COPD patients’ stories of breathlessness: ‘It’s scary/when you can’t breathe’. 14(6), 760–778. 10.1177/1049732304265973.

Braun V and Clarke V (2006) Using thematic analysis in psychology. 3(2), 77–101. 10.1191/1478088706qp063oa.

Braun V and Clarke V (2021) Thematic Analysis: A Practical Guide. SAGE Publications. https://uk.sagepub.com/en-gb/eur/thematic-analysis/book248481 (accessed 2 March 2025)

Christiansen CF, Løkke A, Bregnballe V, Prior TS and Farver-Vestergaard I (2023) COPD-Related Anxiety: A Systematic Review of Patient Perspectives. 18, 1031–1046. 10.2147/COPD.S404701.

Clarke V and Braun V (2017) Thematic analysis. 12(3), 297–298. 10.1080/17439760.2016.1262613.

Creswell JW (2009) Editorial: Mapping the Field of Mixed Methods Research. 3(2), 95–108. 10.1177/1558689808330883.

Farquhar MC, Prevost AT, McCrone P, Brafman-Price B, Bentley A, Higginson IJ, Todd CJ and Booth S (2016) The clinical and cost effectiveness of a Breathlessness Intervention Service for patients with advanced non-malignant disease and their informal carers: mixed findings of a mixed method randomised controlled trial. 17, 185. 10.1186/s13063-016-1304-6.

Global Strategy for Prevention, Diagnosis and Management of COPD: 2025 Report (2025). Retrieved from https://goldcopd.org/2025-gold-report/

Hearn J and Higginson IJ (1999) Development and validation of a core outcome measure for palliative care: the palliative care outcome scale. Palliative Care Core Audit Project Advisory Group. 8(4), 219–227. 10.1136/qshc.8.4.219.

Higginson IJ, Bausewein C, Reilly CC, Gao W, Gysels M, Dzingina M, McCrone P, Booth S, Jolley CJ and Moxham J (2014) An integrated palliative and respiratory care service for patients with advanced disease and refractory breathlessness: a randomised controlled trial. 2(12), 979–987. 10.1016/S2213-2600(14)70226-7.

Maher TM (2024) Interstitial Lung Disease: A Review. 331(19), 1655–1665. 10.1001/jama.2024.3669.

Momtazmanesh S, Moghaddam SS, Ghamari S-H, et al. (2023) Global burden of chronic respiratory diseases and risk factors, 1990–2019: an update from the Global Burden of Disease Study 2019. 59. 10.1016/j.eclinm.2023.101936.

Müllerová H, Lu C, Li H and Tabberer M (2014) Prevalence and Burden of Breathlessness in Patients with Chronic Obstructive Pulmonary Disease Managed in Primary Care. 9(1), e85540. 10.1371/journal.pone.0085540.

Powell B (2014) Managing breathlessness in advanced disease. 14(3), 308–311. 10.7861/clinmedicine.14-3-308.

Rzadkiewicz M, Bråtas O and Espnes GA (2016) What else should we know about experiencing COPD? A narrative review in search of patients’ psychological burden alleviation. 11, 2295–2304. 10.2147/COPD.S109700.

Shenton AK (2004) Strategies for ensuring trustworthiness in qualitative research projects. 22(2), 63–75. 10.3233/EFI-2004-22201.

Silverman D (2013) Doing Qualitative Research: A Practical Handbook. SAGE Publications. (accessed 2 March 2025)

Willgoss TG and Yohannes AM (2013) Anxiety disorders in patients with COPD: a systematic review. 58(5), 858–866. 10.4187/respcare.01862.

Yohannes AM (2020) Depression and anxiety in patients with interstitial lung disease. 14(9), 859–862. 10.1080/17476348.2020.1776118.

